# A Comparative Study of Explainability Methods for Whole Slide Classification of Lymph Node Metastases using Vision Transformers

**DOI:** 10.1101/2024.05.07.24306815

**Authors:** Jens Rahnfeld, Mehdi Naouar, Gabriel Kalweit, Joschka Boedecker, Estelle Dubruc, Maria Kalweit

**Author notes:** these authors contributed equally to this work (Jens Rahnfeld, Mehdi Naouar, Gabriel Kalweit).

## Abstract

Recent advancements in deep learning (DL), such as transformer networks, have shown promise in enhancing the performance of medical image analysis. In pathology, automated whole slide imaging (WSI) has transformed clinical workflows by streamlining routine tasks and diagnostic and prognostic support. However, the lack of transparency of DL models, often described as “black boxes”, poses a significant barrier to their clinical adoption. This necessitates the use of explainable AI methods (xAI) to clarify the decision-making processes of the models. Heatmaps can provide clinicians visual representations that highlight areas of interest or concern for the prediction of the specific model. Generating them from deep neural networks, especially from vision transformers, is non-trivial, as typically their self-attention mechanisms can lead to overconfident artifacts. The aim of this work is to evaluate current xAI methods for transformer models in order to assess which yields the best heatmaps in the histopathological context. Our study undertakes a comparative analysis for classifying a publicly available dataset comprising of N=400 WSIs of lymph node metastases of breast cancer patients. Our findings indicate that heatmaps calculated from Attention Rollout and Integrated Gradients are limited by artifacts and in quantitative performance. In contrast, removal-based methods like RISE and ViT-Shapley exhibit better qualitative attribution maps, showing better results in the well-known interpretability metrics for insertion and deletion. In addition, ViT-Shapley shows faster runtime and the most promising, reliable and practical heatmaps. Incorporating the heatmaps generated from approximate Shapley values in pathology reports could help to integrate xAI in the clinical workflow and increase trust in a scalable manner.

## Introduction

Recent advances in deep learning (DL)^1–3^ have enhanced medical image analysis, including the classification of gigapixel whole slide images (WSIs) in histopathology^4,5^. The impact of automated WSI examination is immense in terms of resources, quality and clinical decision support, leading to a strong current to integrate these technologies into clinical workflows. As a result, a growing number of AI models for histopathological image analysis and in general are already FDA approved^6^. The ability of deep neural networks to process and analyze large amounts of data can assist human pathologists in inspecting WSIs to enhance clinical workflows and accuracy, potentially detecting patterns within WSIs that may not be immediately apparent to the human eye and facilitating a more comprehensive assessment of the slides^7,8^. However, even though current sophisticated systems yield good performance in quantitative evaluations, the integration of DL models into clinical practice is hampered by the complexity and lack of transparency of these *black box* algorithms. It is not obvious for humans, *how* DL models derive their conclusions, which poses significant challenges for clinical acceptance^9^. Practitioners require not only accurate predictions, but also an understanding of the reasoning behind these predictions to ensure trust and the effective use of such technology. This necessitates the implementation of explainability methods^10^, which aim to make the decision-making processes of DL models transparent and understandable to human users. To estimate the most influential parts of an input image for a model, one can rely on several measures, such as gradient, model attention, or other auxiliary measures derived from the sensitivity of a model to omitted inputs through a *surrogate model*. Regardless of how the influence is measured, the attribution measure can be normalized to arrive at the common visualization of *red* and *blue* coloured heatmaps. The more *red* an area of an input image is, the more important it is considered to be. Ensuring explainability is crucial for validating diagnostic recommendations of the model, facilitating the identification of potential biases, and ensuring that the models’ decisions are based on clinically relevant features within the WSIs. It is key in understanding when to *use* and *not to use* an approximate, data-driven model. Transformer models^11^ have set new benchmarks and a new standard in the domain of natural language processing and are increasingly being applied to image analysis^12^, including medical image classification^13^. These models, which are mainly characterized by their self-attention mechanisms, offer significant advancements in capturing long-range dependencies within images. The application of transformer models to the classification of WSIs in the field of oncology promises improvements in the qualitative accuracy, as well as potentially lowering temporal requirements^14^. However, scaling explainability to transformer models is non-trivial^15^ due to the complexity inherent in their architecture and necessitates adaptation of existing methods to ensure that the decision-making processes remain interpretable to clinicians.

Our study focuses on the comparative analysis of state-of-the-art explainability methods applied to transformer models for the classification of WSIs in lymph node metastases among women with breast cancer. We evaluate an extension of the experimental framework by Covert et al. (2023)^15^, which constituted an investigation on the use of approximate Shapley values for vision transformers, and benchmark ViT-Shapley^15^, Attention Rollout^16^ with residuals, Integrated Gradients^17^ and RISE^18^ with varying sampling strategies in this domain. For evaluation, we use the publicly available *CAMELYON16* dataset^19^ consisting of WSIs of lymph node sections from breast cancer patients. This dataset is a standardized benchmark, which includes a comprehensive representation of the histopathological variability present in breast cancer metastases and provides a robust foundation for evaluating the performance and interpretability of explainability methods in a clinically relevant context. On the basis of the classifier proposed by Naouar et al.^20^, we show that meaningful attribution maps can be obtained for gigapixel WSIs using state-of-the-art attribution methods and compare their respective performance and resource requirements through qualitative and quantitative assessment. Our results suggest that the trustworthiness and transparency of expressive transformer-based black-box classifiers for sentinel lymph node sections from breast cancer patients can be enhanced by current attribution methods. To the best of our knowledge, this is the first comprehensive study and comparison of the scalability of explainability methods in this area.

## Results

The study utilized the CAMELYON16 dataset, consisting of N=400 hematoxylin and eosin (H&E) stained WSIs of sections of sentinel lymph nodes. As classifier, we used a Vision Transformer model trained on 20× magnification^20^, which had an AUROC of 96.3 ± 0.1 and an MCC of 55.16 ± 1.11. Additionally, it had a precision of 48.02 ± 1.73, a recall of 80.23 ± 0.81 and a specificity of 98.72 ± 0.14. On this basis, we assessed the capabilities of several explainability methods for this classifier: Attention Rollout with and without residuals, Integrated Gradients, RISE with both binominal and uniform mask sampling, and Shapley values. Below, we provide an in-depth explanation of how these different methods work.

### Qualitative Assessment

The qualitative assessment was conducted by an expert pathologist. The aim was to assess the logical coherence of the heatmaps identified by the examined explainability methods, which ought to highlight regions determined to be important for the decision of the trained Vision transformer classifiers. Visual analysis showed (cf. Figures 1 to 3), that – despite highlighting some of the tumor cells – Attention Rollout methods tend to highlight overconfident artifacts, emphasizing non-informative regions, such as the background. The incorporation of residuals intensified these effects. Integrated Gradients primarily focused on a subset of individual tumor cells. In contrast, both RISE and Shapley methods generally highlighted areas extensively covered by tumor cells. While RISE distributes more importance and variance to background regions, Shapely shows the most concised heatmaps with strongest focus on the complete set of visible tumor cells.

**Figure 1.**
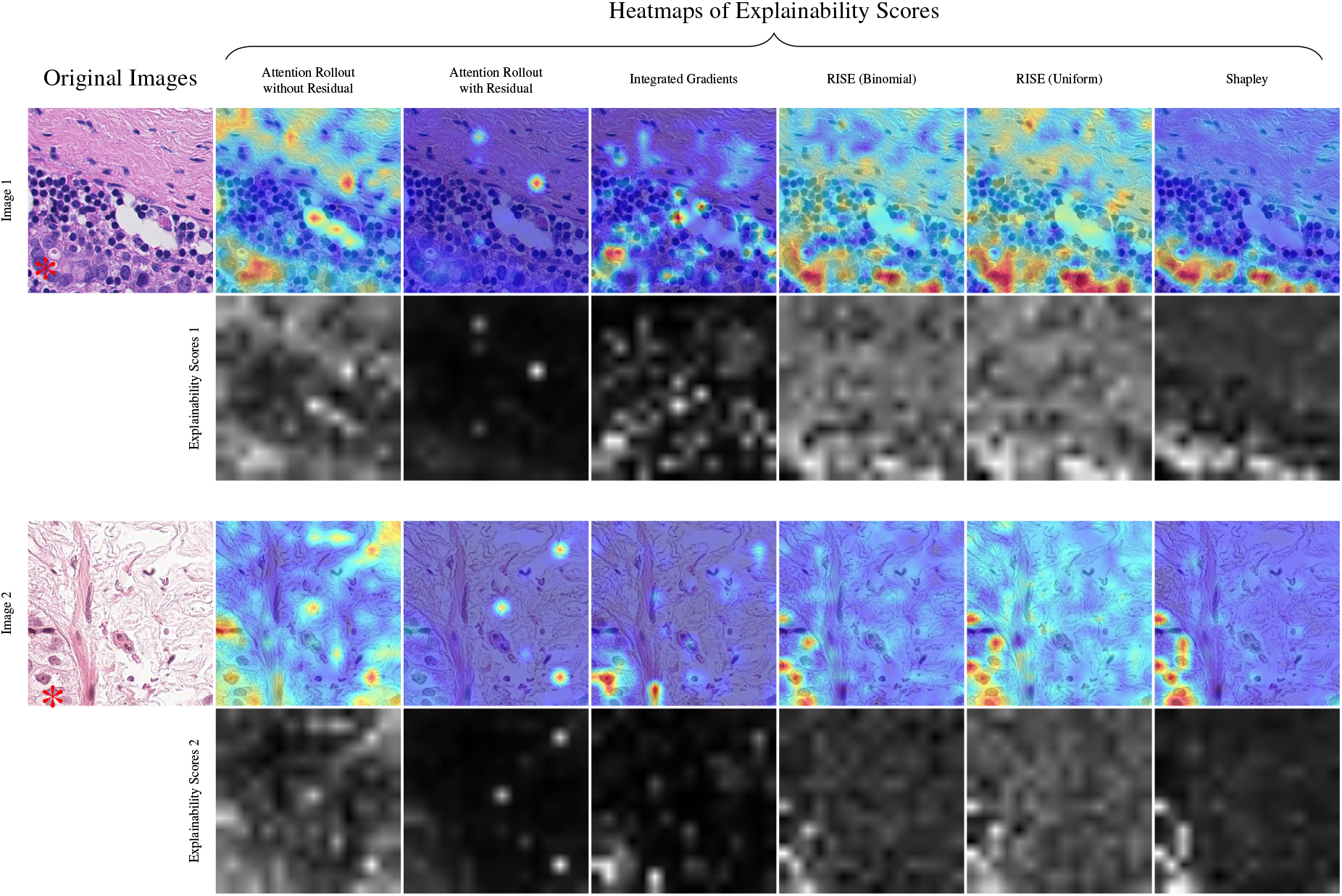
Qualitative Examples for different heatmaps of explainability scores for a vision transformer from different explaination methods on patches from WSI test_110 and test_052 of the *Camelyon16 dataset*. Original images are shown on the left, with metastatic cells on the lower part. Tumor tissue is marked with *. The first row shows each method’s explainability scores as heatmaps blended over the image, and the second row the corresponding raw explainability scores inferred from the trained vision transformer. Shapley (on the right) shows the clearest distinction between tumoral and non-tumoral tissue.

**Figure 2.**
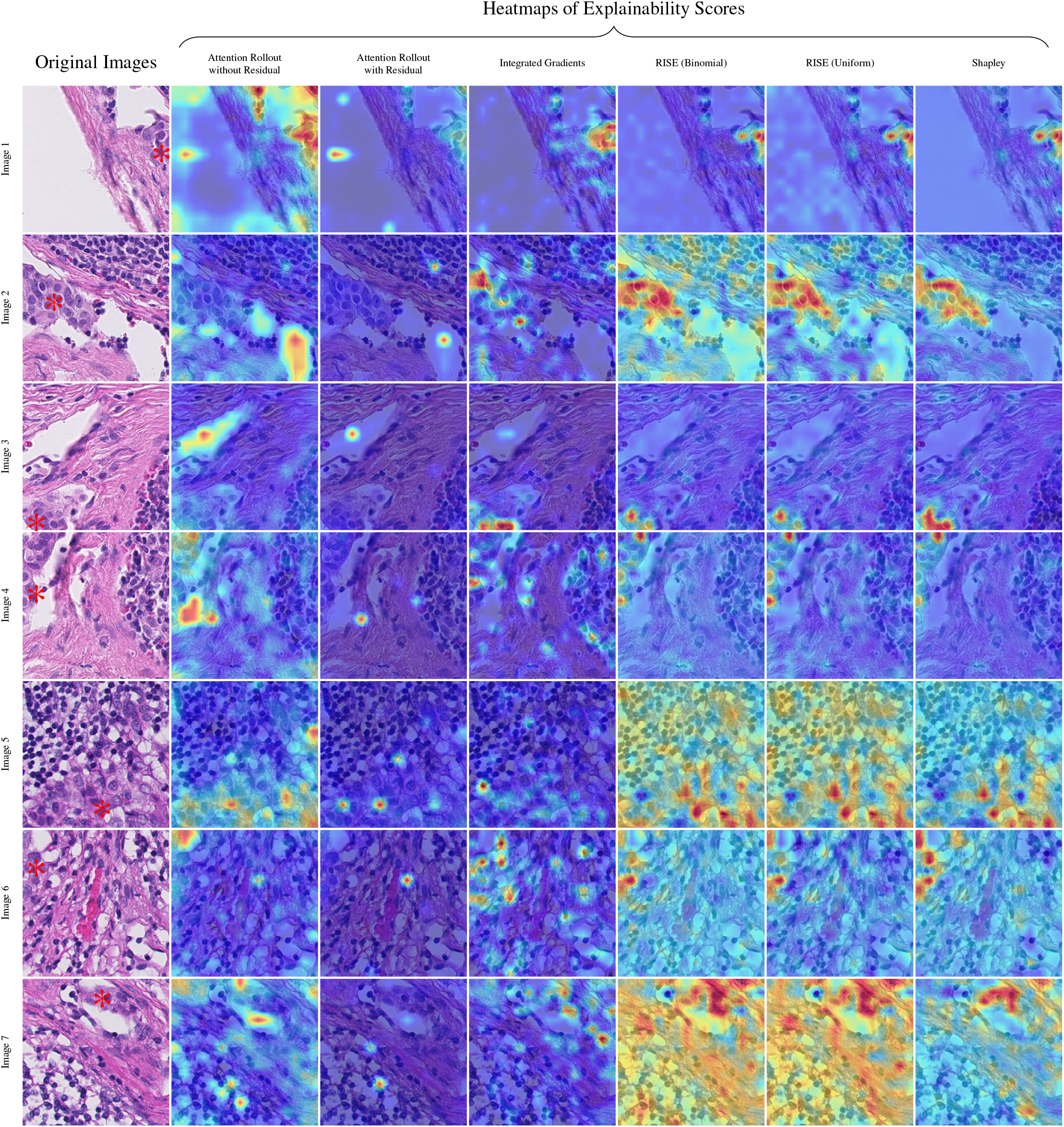
Further qualitative examples for patches from test_004 and test_008 of the *CAMELYON16* dataset. Tumor tissue is marked with * in the original image. Each method’s explainability scores are shown as heatmaps blended over the image.

**Figure 3.**
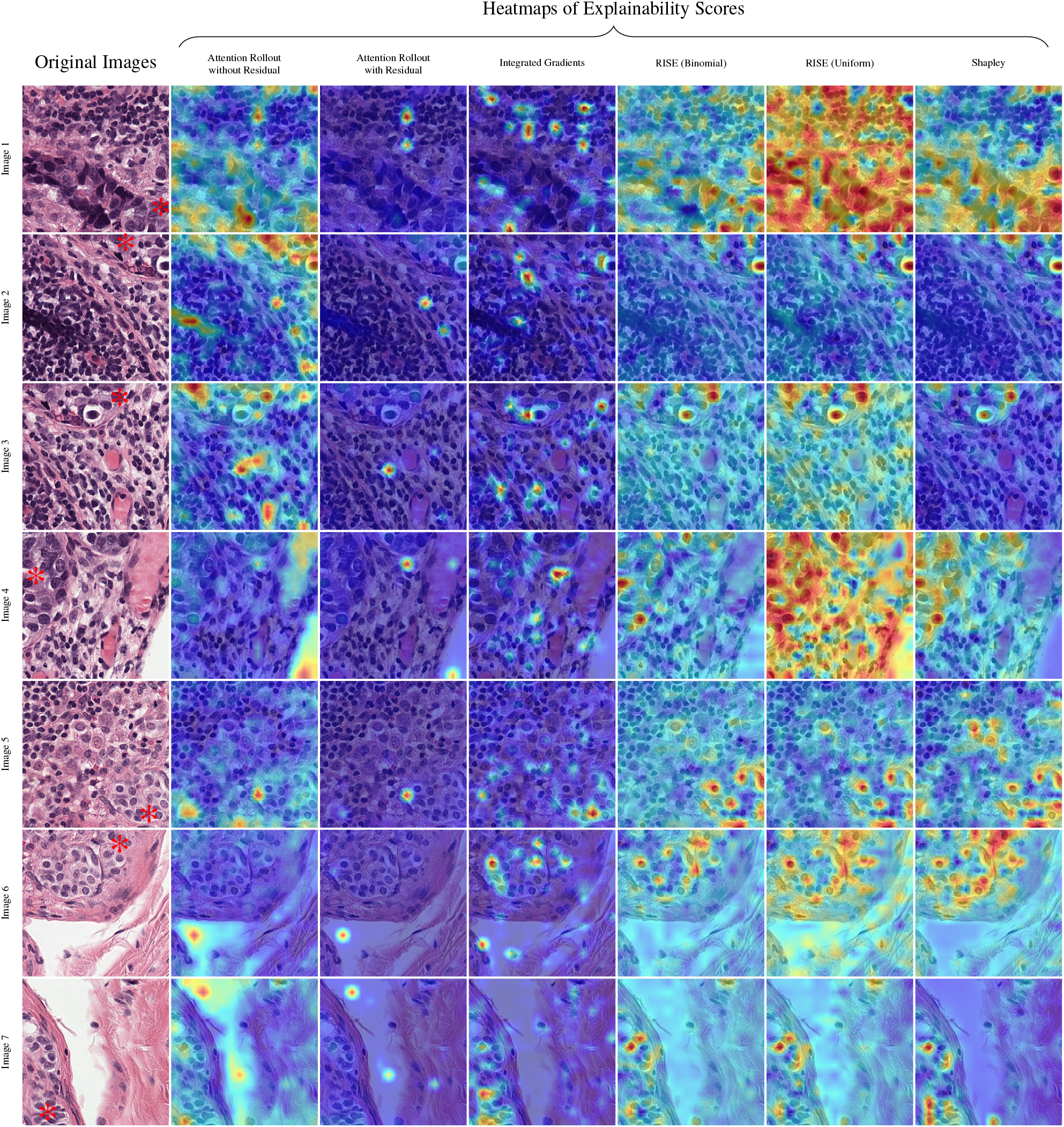
Further qualitative examples for patches from test_010 and test_013 of the *CAMELYON16* dataset. For every image, each method’s explainability scores as heatmaps are blended over the image. Tumor tissue is marked with *.

### Quantitative Evaluation

For a quantitative evaluation, we focus on the widely used *Insertion* and *Deletion* metrics^18^, for which we generate predictions while inserting or removing features in the order of most to least importance and evaluate the area under the curve of prediction probabilities^1^. Our analysis reveals consistent patterns across methods, as depicted in Figures 4a to 4d and detailed in Table 1. Initial analyses confirmed that all evaluated methods outperformed random attribution maps significantly (^****^ *p* < 1.65 · 10^−36^; Welch’s t-test). Attention Rollout, enhanced with residuals in the attention matrix, exhibited superior performance in both metrics. Integrated Gradients achieved comparable Insertion scores to Attention Rollout but fell short in Deletion metrics (^*^ *p* < 0.0465; Welch’s t-test). Notably, removal-based methods demonstrated significantly superior performance (^***^ *p* < 4.61 · 10^−4^; Welch’s t-test). RISE, with binomially and uniformly distributed mask cardinalities, showed marginally higher AUC for Insertion (^****^ *p* < 1.33 · 10^−7^; Welch’s t-test), while Shapley’s lower Deletion AUC indicated better performance compared to both RISE variants (^****^ *p* < 7.13 · 10^−47^; Welch’s t-test). Remarkably, RISE performed better with binomial distribution of mask cardinality (^****^ *p* < 6.87 · 10^−8^; Welch’s t-test). Furthermore, averaging scores over WSIs generally resulted in lower Deletion scores than averaging across individual patches, highlighting the importance of aggregation method on performance evaluation.

**Table 1.** Comparison of methods based on Mean over all individual patch AUCs and Mean over per whole slide image averaged AUC. For *Insertion*, higher is better (↑); for *Deletion*, lower is better (↓).

| Method | Mean over WSIs |  | Mean over patches |  |
| --- | --- | --- | --- | --- |
| | Insertion ↑ | Deletion ↓ | Insertion ↑ | Deletion ↓ |
| Attention Rollout (without residual) | 0.8170 | 0.5297 | 0.8449 | 0.6004 |
| Attention Rollout (with residual) | 0.8558 | 0.4412 | 0.8791 | 0.5318 |
| Integrated Gradients | 0.8559 | 0.6457 | 0.8766 | 0.7117 |
| Random | 0.7740 | 0.7685 | 0.8091 | 0.8034 |
| RISE (binomial) | **0.9834** | 0.2291 | **0.9838** | 0.2947 |
| RISE (uniform) | 0.9794 | 0.2626 | 0.9799 | 0.3378 |
| Shapley | 0.9740 | **0.1866** | 0.9751 | **0.2511** |

**Figure 4.**
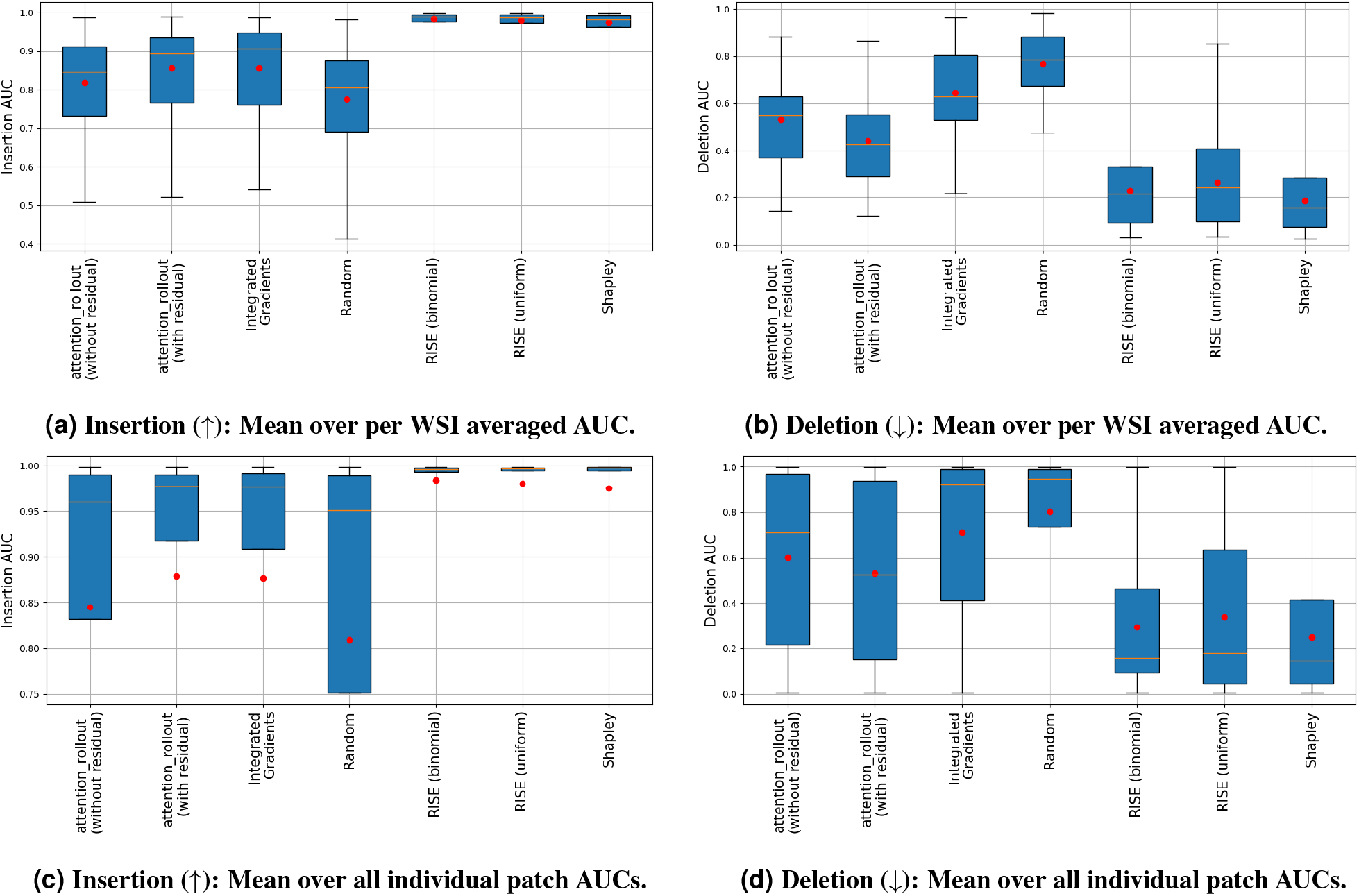
Mean Insertion per WSI (a) and over all individual patches (c) and mean Deletion per WSI (b) and over all individual patches (d). Each mean Insertion (a) and Deletion (b) AUC of a whole slide image represents a sample. (c) Each individual patch’ Insertion (c) and Deletion (d) AUC represents a sample. Red dot is the mean within the populations.

#### Computational Complexity

We benchmarked RISE and ViT-Shapley on a single NVIDIA GeForce RTX 2080 Ti with 20 CPU Cores (Intel(R) Xeon(R) Silver 4210 CPU @ 2.20GHz). The surrogate model’s underlying backbone is a ViT-Small with a patch size of 16 imported from the ^2^timm library (vit_small_patch16_224). A comparison for compute and memory requirements of RISE and ViT-Shapley can be found in Figure 5. Shapley is by order of magnitudes faster with a runtime of 1.547 seconds for a single patch while RISE takes 4.978 seconds when fully parallelizing the sampling. With our hardware constraints, we found 10 masks per GPU and a batch size of 96 for RISE to be a good trade-off between *processing time of a single batch* vs. *throughput* which required 317 seconds. In our test set, the median number of tumor patches occuring in a WSI was 283 patches. Thus, processing such a WSI would take about 16 minutes. On the other hand, for Shapley, we could fit a batch size of 32 onto our GPU leading to a total processing time of about 14 seconds.

**Figure 5.**
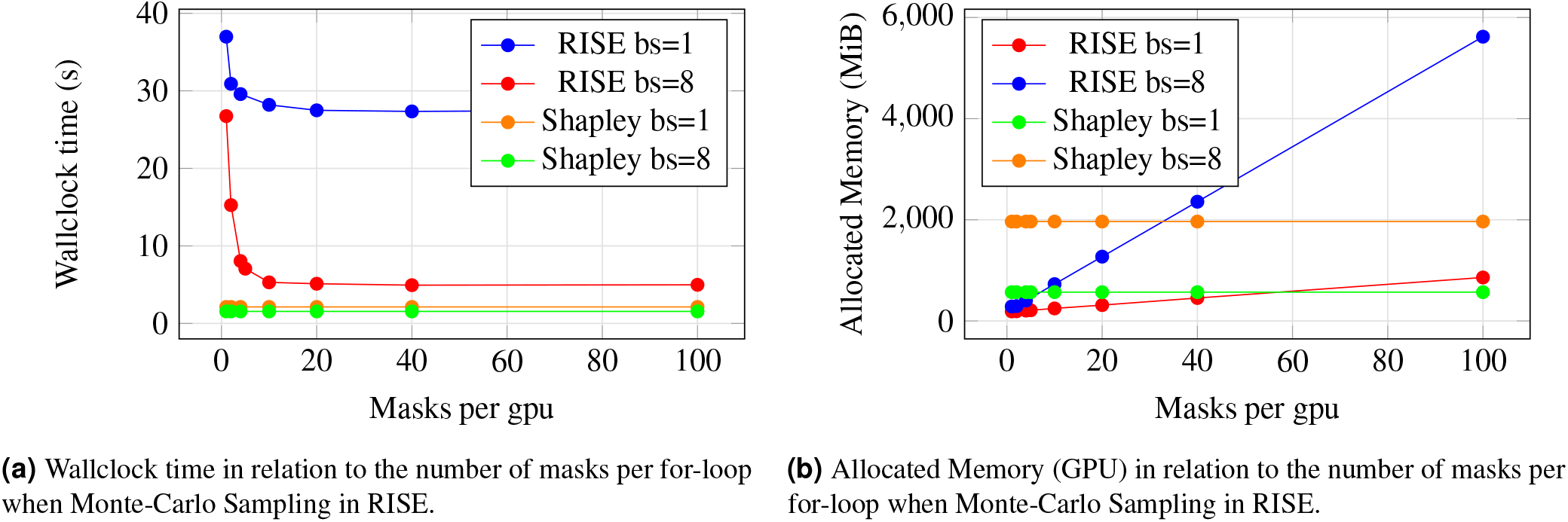
Comparative analysis of wallclock time and allocated GPU memory in RISE.

## Discussion

When assessing the effectiveness and usefulness of explainability methods in machine learning classification, it is first important to set clear expectations. Effective explainability methods should highlight the factors that a classifier relies on to make its decisions, which may *match* human expectations or reveal *biases*, with both being equally important. Interpreting the results of explainability methods is therefore a complex task. Nevertheless, if a classifier demonstrates high quantitative performance in distinguishing between classes like *tumor* or *non-tumor*, it is reasonable to expect that it has identified significant features of those classes – and thus these features should be reflected by the explainability method and should be in line with the expectations of human experts for *obvious* cases. On the other hand, if a classifier tends to overfit to certain meaningless patterns in the training data, this should be a result of the explainability method just as well. For instance, our classifier was trained to distinguish between lymph node metastasis and normal tissue. It could be the case, that it learned from other features (not just the tumor cells, but from surrounding tissue). Hence, we have to distinguish between useless artifacts and important features.

The observed artifacts in Attention Rollout, characterized by random dots of high perceived importance, negatively impact not only the quantitative outcomes but also the interpretability for healthcare practitioners. These artifacts render the attribution maps confusing, potentially obscuring meaningful insights. In contrast, Integrated Gradients presents a more visually coherent approach by highlighting individual tumor cells. However, it does not perform as well quantitatively, particularly in comparison to removal-based methods. This discrepancy suggests that, while Integrated Gradients may appear reasonable to the human eye, it may not fully capture the network’s reasoning process. Therefore, underscoring the need for attribution metrics that can bypass human biases to accurately evaluate the quality of attribution maps. Nevertheless, relying solely on Insertion and Deletion metrics introduces its own form of inductive bias. Firstly, the concept of “removing information” as a means to assess importance is an artificial construct that may not align with the AI model’s operational principles. Removal-based methods incorporate this concept inherently in their design, in contrast to other methods that do not presuppose any specific mechanism of information assessment. Secondly, Insertion and Deletion ignore the relativity of attribution w.r.t. the ranked information. Thirdly, these metrics do not account for the correlation between patches or the network’s robustness, potentially overlooking the nuanced ways in which AI models derive their classifications. For instance, a network’s focus on a particular tumor cell might be identified as the primary explanation for a classification. However, the model’s ability to maintain accurate classification upon the removal of that cell—by shifting focus to another tumor cell challenges the reliability of such metrics in isolation. This complexity necessitates a balanced approach that combines metric-based evaluations with qualitative assessments of attribution maps or a better scalar measure of attribution.

In this context, RISE and Shapley methods emerge as robust options, offering attribution maps that are both quantitatively and qualitatively trustworthy. While RISE demonstrates competitive outcomes, its utility is tempered by significant inference times, attributed to the Monte-Carlo Sampling process. This limitation poses practical challenges for real-world applications, emphasizing the need for efficient explanation methods. Conversely, Shapley values achieve an optimal balance between trustworthiness and efficiency. After an initial investment in training an explainer network (approximately 3 days), Shapley values enable efficient inference with a single forward pass (approximately 1.547 seconds for a single patch). This efficiency, combined with the method’s reliability, positions Shapley values as a compelling solution for scaling explainability in healthcare AI applications. Thus, our findings advocate for the adoption of Shapley values as a practical and effective approach to understanding and explaining AI model decisions in a clinical context. We therefore consider a combination of vision transformer-based classification, that tend to perform better than established CNN-based methods, and an approximate estimation of corresponding Shapley values to provide the best current solution of automated WSI examination, optimizing for both model performance and explainability. This offers clinicians valuable insight and trustworthiness into the decision making of data-driven visual support systems, potentially enhancing acceptance in clinical workflows.

## Methods

### Dataset Description

The study utilized the CAMELYON16 dataset, consisting of N=400 H&E stained WSIs of sections of sentinel lymph nodes in patients with breast cancer collected in the Radboud University Medical Center Nijmegen and the University Medical Center Utrecht (Netherlands). They come with non-exhaustive expert annotations from pathologists. For training the classifier, we split the WSIs into a training set of 159 tumor-free and 111 tumor slides and a test set of 81 tumor-free and 48 tumor slides (with one WSI being corrupted). We then evaluated the explainability methods on 38 of the 48 tumor slides, omitting the top 10 slides with the most tumor patches to prevent the test set from being highly unbalanced. In total, the resulting test set consisted of 17.343 patches.

### Preprocessing

We first locate tissue inside the WSI and then extract patches of size 256 that are found in the tissue area. We compute an image mask using the *colorization value*^20^ defined as *c*(*r, g, b*) = |*r* − *m*| + |*g* − *m*| + |*b* − *m*|, where 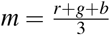. We then apply an adaptive threshold^21^ to separate tissue from background. The colorization value of a pixel is lowest for all shades of grey and is high for all others. The WSIs were split into patches of size 256 on the basis of magnification 20×.

### Pretraining and Model Implementation Details

In order to balance the training data, we perform undersampling on the majority class. We further employ data augmentation via uniform random modulation of brightness, contrast, saturation, hue and flipping. For pre-training of the backbone, we use ImageNet^22^. All models (classifier, surrogate and explainer) are trained on a batch-size of 64 using the Adam optimizer^23^ with *β* = (0.9, 0.999) and *ε* = 10^−8^, and a learning rate of 2.5 · 10^−5^ for the classifier & surrogate and 2.5 · 10^−4^ for the explainer. At the beginning of the training, we warm-start the learning rate linearly followed by a cosine annealing without any restarts. The classifier and surrogate model are trained for 5 epochs while the explainer is trained for 10 epochs. With this setup on one NVIDIA GeForce RTX 2080 Ti, training the classifier takes about 22 hours and surrogate takes about 15 hours. We used two gpus for the explainer which took 2 days & 18 hours. However, there is a bottleneck stemming from loading the whole slide images in our hardware. Without that bottleneck, we expect training time to reduce significantly. The explainer training was limited to 10 epochs mainly due to computational budget constraints.

### Attribution Methods

We compared four different attribution methods: Integrated Gradients, Attention Rollout, Randomized Input Sampling for Explanation and Shapley Values. This subsection introduces aforementioned attribution methods from a theoretical perspective.

#### Integrated Gradients

Integrated Gradients^17^ is an instantiation of Calculus’ fundamental theorem for line integrals which states

##### Theorem 1

*Assume f* : ℝ^*d*^ → ℝ *is a differentiable function and γ* : [*a, b*] → ℝ^*d*^ *a smooth curve. Then*

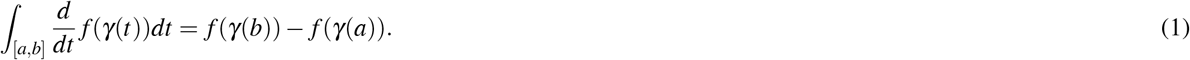

Given a baseline point *x*^*′*^ and target point x, the straight line *γ* : [0, 1] → ℝ^*d*^, *γ*(*t*) = *x*^*′*^ + *t* · (*x* − *x*^*′*^) is a smooth curve and Theorem 1 simplifies to

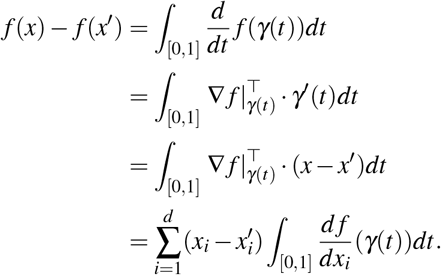

Every summand is now interpreted as the contribution of component *i* to transforming a baseline prediction *f* (*x*^*′*^) into the actual prediction *f* (*x*), i.e.

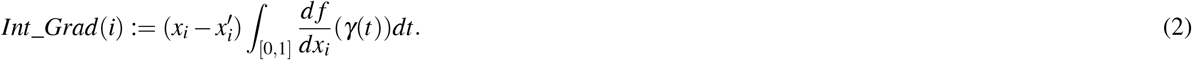

In case of image explanation, the baseline could be a completely masked image *x*^*′*^ = 0 and each image patch would represent a component *i*. This is illustrated in Figure 6. In practice, computing the integrals in Equation (2) exactly is infeasible. Thus,^17^ propose to approximate the integral with a sample size that roughly satisfies the completeness axiom

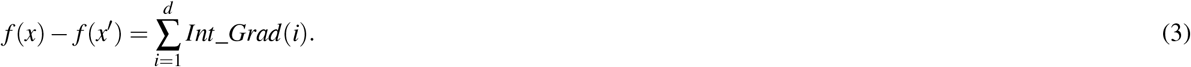

**Figure 6.**
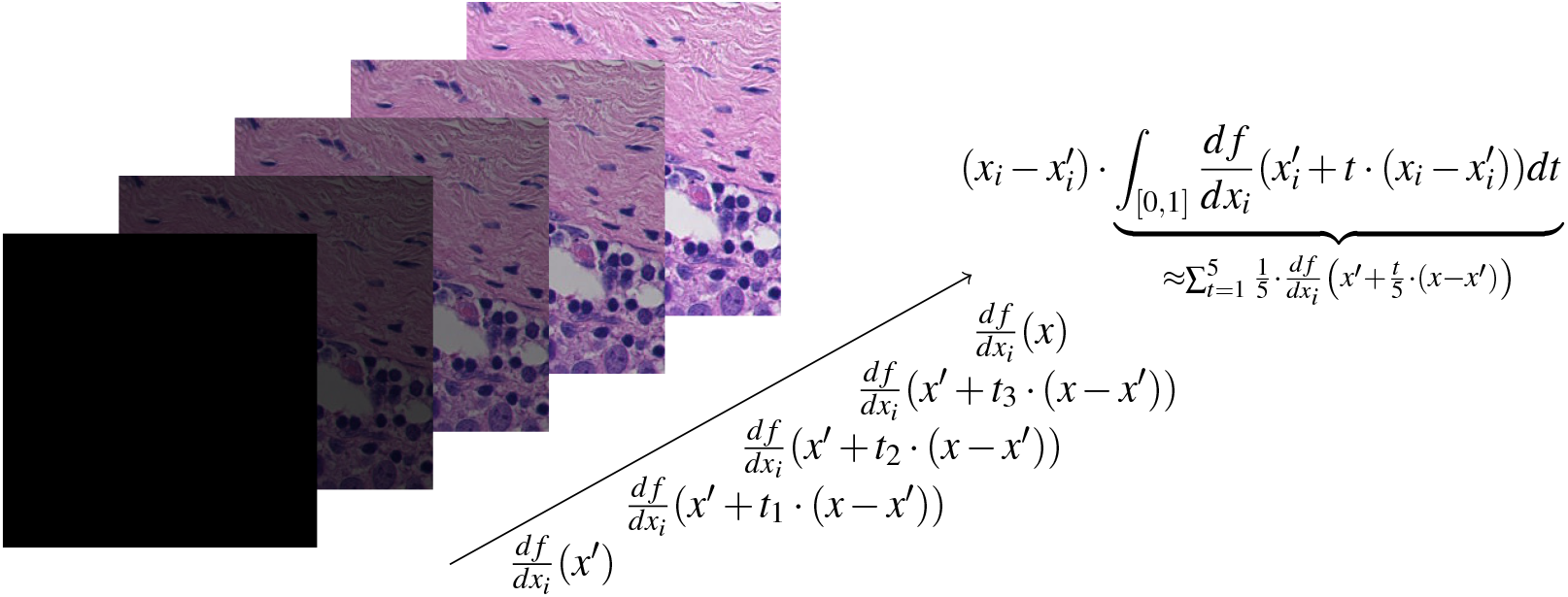
Integrated Gradients for *i*-th component. The line integral is approximated using a Riemann sum with *n* = 5 steps

#### Attention Rollout

A very natural approach to explaining Vision Transformers^24^ is to investigate its attention mechanism^25^. During a forward pass, in each layer *l* = 1, …, *n*, an attention map *A*_*l*_ ∈ ℝ ^(*d*+1)×(*d*+1)^ is computed where *d* denotes the number of patches. Given the token’s value embeddings 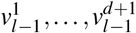 from the previous layer, the next embeddings are retrieved in the following way:

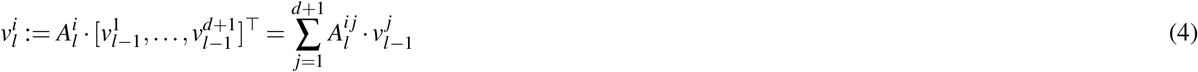

This can be interpreted in the following way: The *i*-th row of *A*_*l*_ represents how much information is retrieved from each of the previous token’s embedding. A visualization of this information flow as bi-partite graph is given in Figure 7. Based on this observation, one could use the last layer’s attention weights pointing to the class token in order to explain which token contributes how much to the Vision Transformer’s prediction. However, it is unclear to which degree the embeddings of the last layer can be associated with the original input tokens. Instead,^16^ propose to join these *l* bi-partite graphs respectively in their shared nodes as illustrated in Figure 8. Formally, we obtain a graph *G* = (*V, E*) with

**Figure 7.**
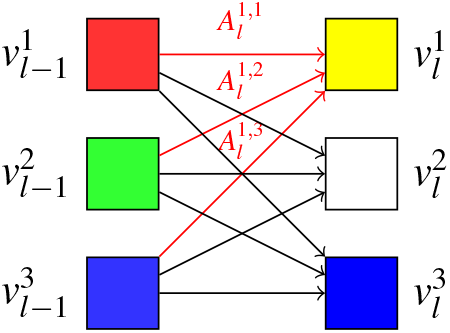
3 × 3 Attention Map interpreted as bi-partite Graph. The edge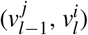 ‘s weights corresponds to the corresponding value in the attention map. The yellow embedding results from retrieving half red, half yellow and zero blue.

**Figure 8.**
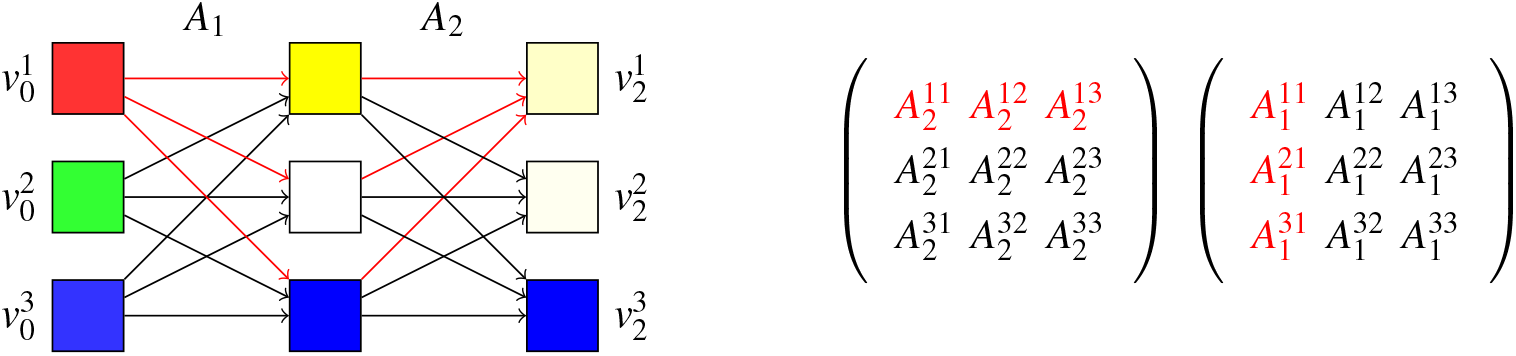
3 × 3 Attention Flow, 2 layers. There are 3 paths (marked in red) representing information flow from 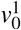 to 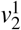. The total amount of information flowing from source to target node is obtained through vector-vector multiplication of the red marked vectors.

- nodes 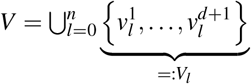
- edges 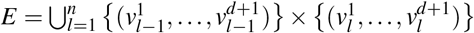
- edge weights 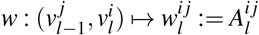

Each path *p* ∈ *V*_0_ ×··· *×V*_*n*_ starting at node 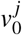 and ending in node 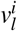 represents information flow from token *j* to token *i* where the amount of information is measured by the product of all traversed edge weights

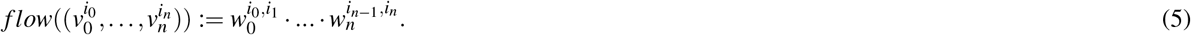

Suppose the first token denotes the class token, i.e. *cls* := 1. Attention Rollout quantifies the importance of token *i* as the sum of all flows from token *i* to the class token

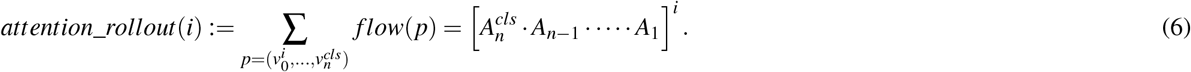

While this tracks flow between attention modules, a Vision Transformer usually applies a skip connection after the attention module, i.e.

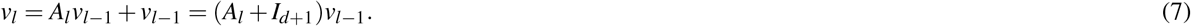

Thus,^16^ extend Equation (6) by adding the identity matrix to each attention map *A*_*l*_ and re-normalizing them row-wise. We denote this as Attention Rollout with residuals.

#### Randomized Input Sampling for Explanation (RISE)

A classifier may not necessarily be based on Vision Transformer rendering Attention Rollout unapplicable. This motivates the class of black-box attribution methods which determine attribution by measuring fluctuations in the target class prediction *f* under image perturbation. Random Input Sampling for Explanation (RISE)^18^ deems an image region / patch important if it contains enough information for the classifier to preserve its high class probability under removal of other regions / patches. In other words, a region / patch *i* is important if *f* is high in expectation over arbitrary masking of other regions, i.e.

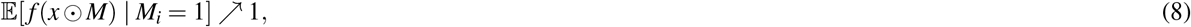

where *x* is the input image and *M* ∼ *P*_*M*_ a random variable mapping to {0, 1}^*d*^. We can re-write Equation (8) in the following way

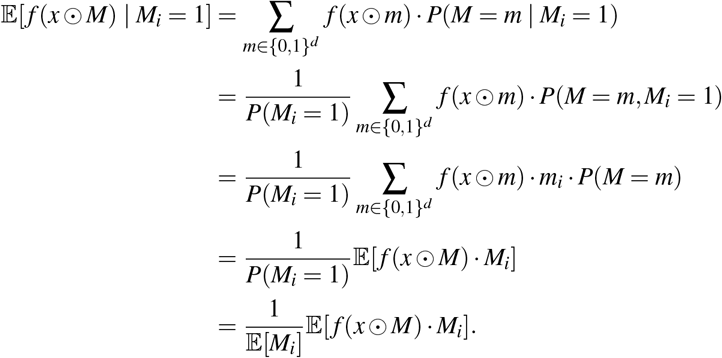

Note that 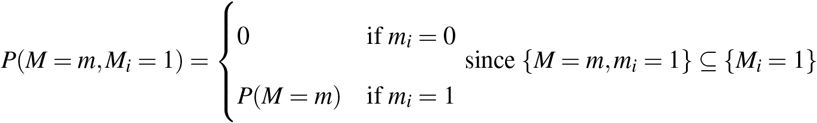. Thus, re-writing Equation (8) for every patch i, we obtain the following expectation in matrix notation

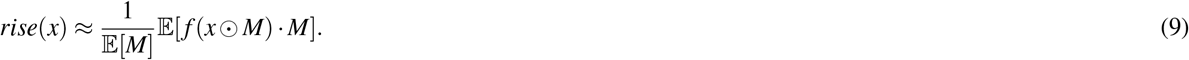

which is approximated using Monte Carlo Sampling. The Monte Carlo Sampling procedure is illustrated in Figure 9.

**Figure 9.**
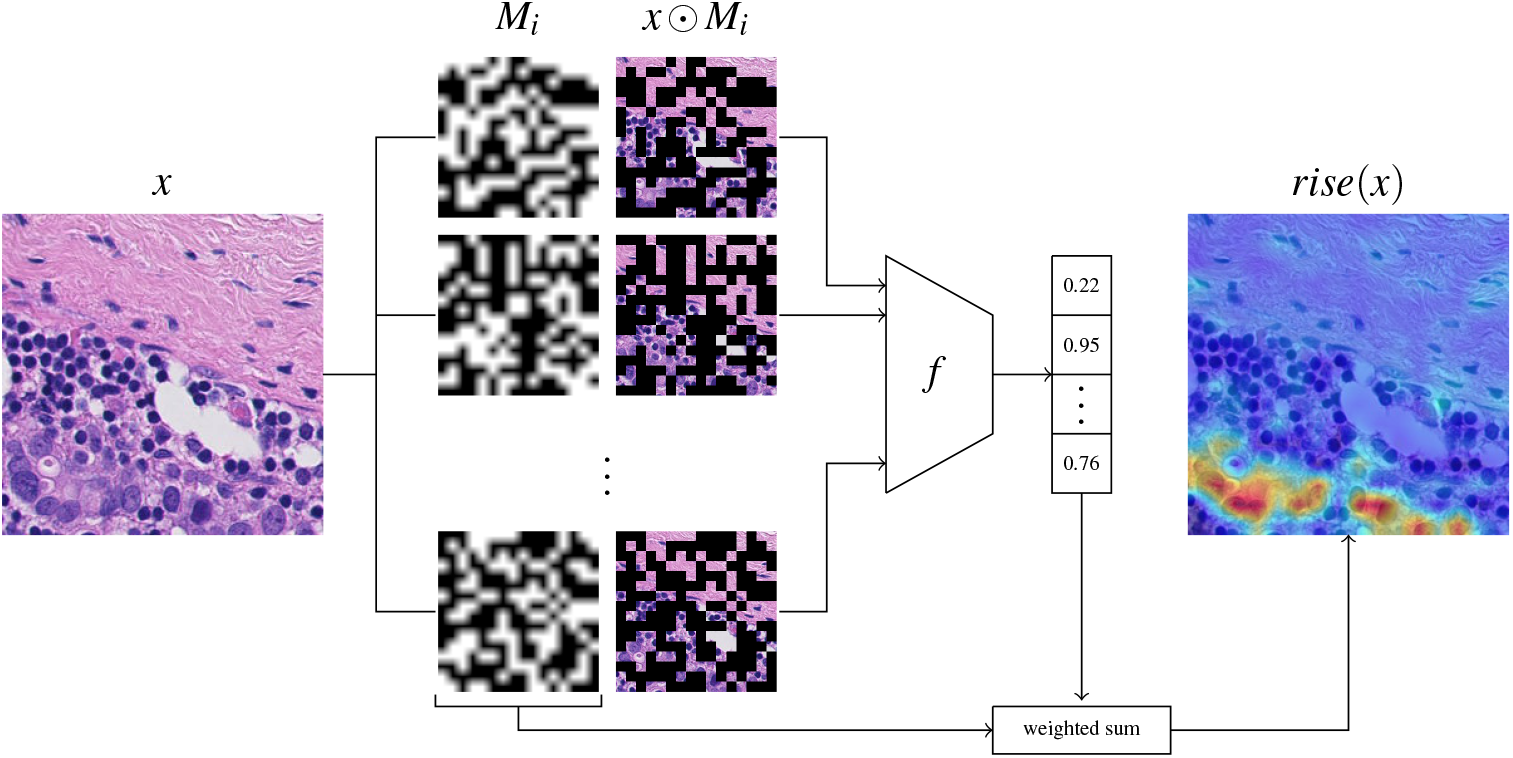
Randomized Input Sampling for Explanation (RISE). Monte Carle Sampling with n masks *M*_1_, …, *M*_*n*_.

#### Shapley Values

Stemming from Game Theory, Shapley Values^26^ have been introduced for *d*-player cooperative games to determine how much each player contributes to the group’s overall payout. A *d*-player cooperative game is defined via a set function *v* : {0, 1}^*d*^ → ℝ satisfying *v*(0) = 0. Evaluating *v*(*m*) amounts to grouping a subset of players in a coalition (*m*_*i*_ = 1 ⇔ player *i* is included), letting this coalition play out the game and obtaining their payout. The idea is now to compare for every coalition *m* ∈ {0, 1}^*d*^ its payout

a. with player *i*: *v*(*m* + *e*_*i*_) vs.
b. without player *i*: *v*(*m*).

Player *i*’s Shapley Value *ϕ* _*i*_ ∈ ℝ is then defined as

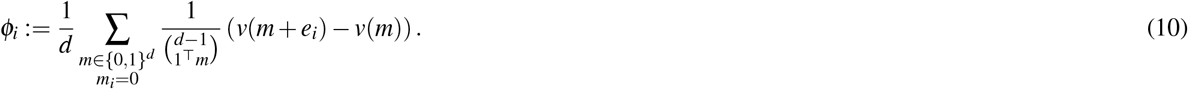

Note that for fixed coalition size *k* = 1^T^*m* we can draw 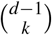 different coalitions out of the player pool {1,…, *d*} \ {*i*} without player *i*. In other words, Equation (10) balances out term’s weights such that coalitions of size *k* have the same influence on the Shapley Value as coalitions of size *k*^*′*^ ≠ *k*. Unfortunately, computing Equation (10) is very expensive as it requires 2^*d*^ queries of *v*.^27,28^ derived an alternative characterization as least squares solution of the following optimization problem

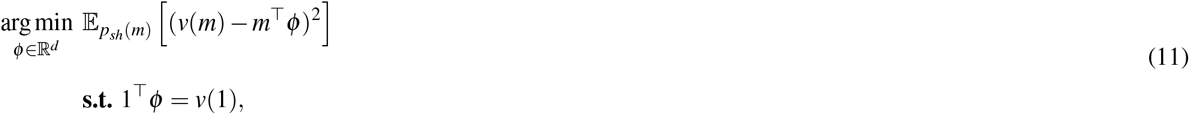

where

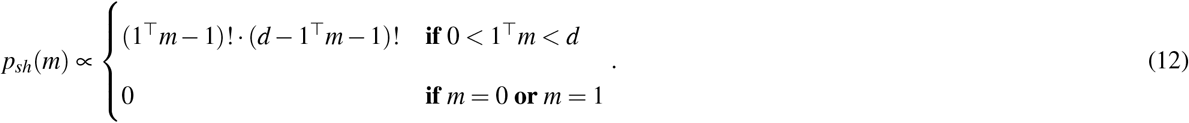

Equation (11) can now be solved using a projected stochastic gradient descent approach. To this end,^28^ introduce additive efficient normalization

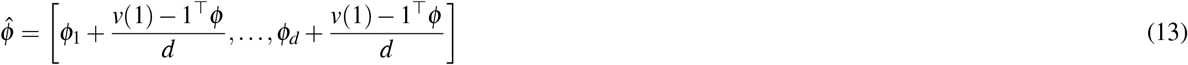

which projects back a candidate solution *ϕ* to the feasible space since

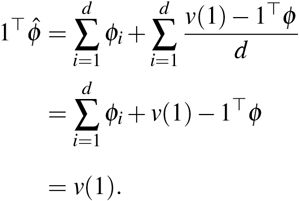

Furthermore,^28^ show that the resulting optimum does indeed solve Equation (11). Bridging the gap to images, for any image *x* the goal is to explain a classifier 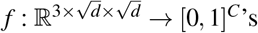 probability of a target class *y*. Thus, every patch of the image represents a player. The game *v*_*xy*_ : {0, 1}^*d*^ → ℝ to be played amounts to evaluating the classifier on the image where the corresponding patches are masked out. Concretely,

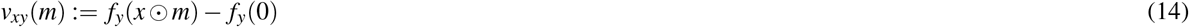

where *f*_*y*_(0) denotes the baseline prediction, i.e. evaluating on the black image. Subtracting the baseline prediction is required to satisfy *v*_*xy*_(0) = 0. Within the realm of Vision Transformer’s,^15^ introduce an explainer network 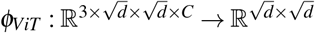 which learns to predict the Shapley Values regarding all classes *y* ∈ *C* for any image *x*. This is done by minimizing the loss

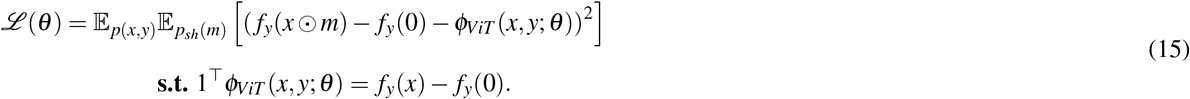

The explainer model *ϕ* _*ViT*_ applies additive normalization as outlined in Equation (13) after each forward pass, even during inference. So by default, the side constraint in Equation (15) is satisfied reducing the constrained problem to a plain minimization problem which can be tackled using using standard optimizers such as SGD, Adam^23^, AdamW^29^, etc.

Usually, classifiers are not trained on images with missing patch information. Thus, evaluating the classifier on masked images requires marginalizing over the missing patch information, i.e. computing

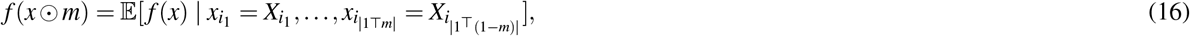

where 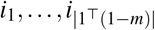 index the missing patches when masking with *m* ∈ {0, 1}^*d*^, i.e.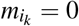. To tackle this problem,^15^ additionally train a surrogate model *g* : 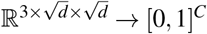 by minimizing

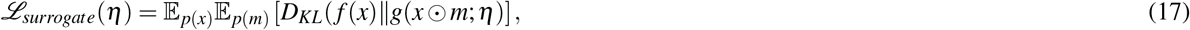

where

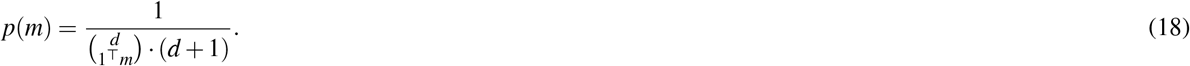

Intuitively, sampling from Equation (18) can be thought of as uniformly sampling a cardinality *k* ∈ {0,…, *d*} followed by drawing a mask *m* out of all 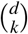 possible masks with cardinality *k*. The surrogate model *g* then replaces *f* in Equation (15).

### Attribution Metrics

Human inspection of qualitative examples is inherently biased by our intuition about “logically” appearing attribution maps. In that sense, neural networks may have a completely different way of perception and reasoning. This motivates the need for more mathematically grounded and objective metrics.^15^ rely on 4 such metrics: Insertion, Deletion^18^, Faithfulness^30^ and Sensitivity-n^31^.

#### Insertion and Deletion

When inspecting an image, humans usually only need a few select image patches to infer the corresponding object’s class. So, starting from a black image, if we were given the first few important patches step-by-step, our confidence in predicting the target class would quickly rise. Conversely, if given the wrong patches, i.e. with no useful information, we would remain rather unconfident in predicting the target class for a longer period of time. As a result, suppose we were to plot our confidence score *f*, i.e. predicted class probability, with regards to the number of inserted patches. The more meaningful the first few patches are, the quicker we would expect the curve to converge to a high probability. This is illustrated in Figure 10. How quickly the curve converges to a high probability is then directly linked to the amount of area it covers. Mathematically, given the ranking *rank* : {1,…, *d*} → {1,… *d*} resulting from argsorting the attribution map, **Insertion** measures the area under the curve of the plot

**Figure 10.**
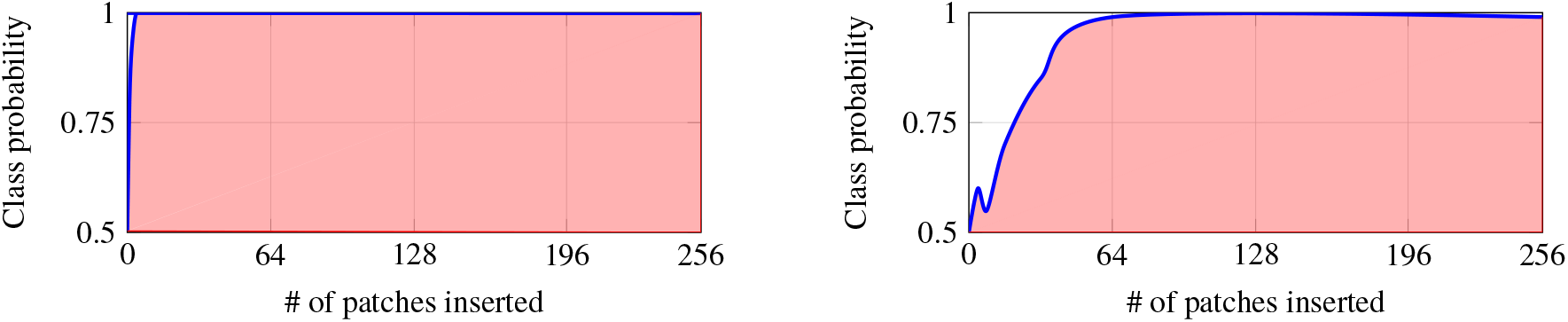
Classifier’s predicted class probability when adding patches step-by-step. Left: Important patches are inserted from the beginning leading to a high area under the curve. Right: Meaningless patches are inserted first. Thus, having a lower area under the curve.

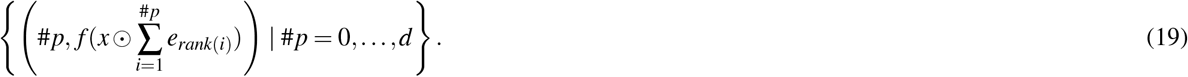

Higher is better. With the same intuition as above described in mind, we could also do this the other way around. That is, start with the whole image and step-by-step remove the patches according to the ranking in a descending order. If the first few patches were the most meaningful, then we would expect the class probability curve to quickly converge down to some low value. In other words, the covered area would be low this time. Mathematically, **Deletion** measures the area under the curve of the plot

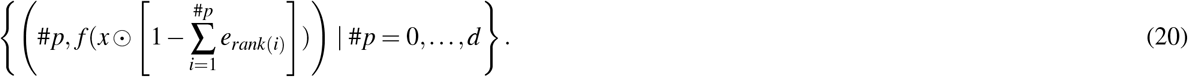

Lower is better.

#### Faithfulness and Sensitivity-n

Ranking-based metrics ignore the concrete values of the attribution map. Sensitivity-n and Faithfulness try to incorporate this additional information by measuring how much the predicted class probability drop under masking out is correlated to the corresponding assigned attribution values. If a drop is high, then we would expect the masked out patches to have contributed significantly, i.e. a high attribution value too.

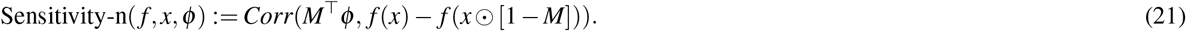

where the random variable *M* ∈ {0, 1}^*d*^ realizes masks of cardinality *n* drawn uniformly, i.e.

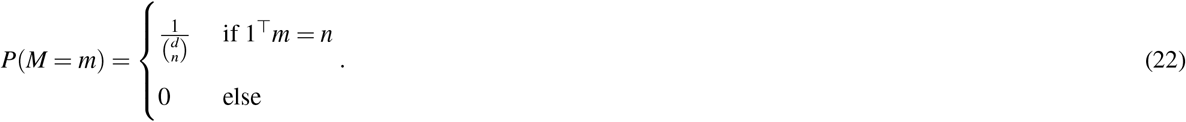

Instead of iterating over a preset of cardinalities *n*, the correlation can also be measured over all masks.

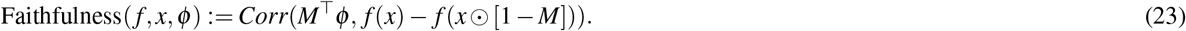

where the random variable *M* ∈ {0, 1}^*d*^ realizes masks drawn from the distribution specified in Equation (18). In both cases, higher is better. Furthermore, computing the expectation exactly is infeasible. Thus, both metrics are approximated through Monte-Carlo sampling.

### Statistical Summary

Table 2 presents a statistical overview of the performance metrics, detailing mean AUC scores averaged per WSI and across all individual patches, respectively. In the table, p-values are shown for individual patch AUCs as sample populations and for per whole slide image averaged AUCs. However, it should be pointed out that each patient’s whole slide image provided a different number of patches. Thus, the samples are not i.i.d.

**Table 2.** Welch’s t-test’ p-values for Insertion and Deletion. Significant values are marked in red.

|  | Attn. Roll.<br>(w/o res) | Attn. Roll.<br>(with res) | integrated<br>gradients | random | rise<br>(binomial) | rise<br>(uniform) | shapley |
| --- | --- | --- | --- | --- | --- | --- | --- |
| Insertion: Mean over per whole slide image averaged AUC |  |  |  |  |  |  |  |
| Attn. Roll.<br>(w/o res) | - | 1.0 | 1.0 | 1.0 | 1.370e-07 | 2.424e-07 | 5.195e-07 |
| Attn. Roll.<br>(with res) |  | - | 1.0 | 4.151e-01 | 3.908e-06 | 7.782e-06 | 1.917e-05 |
| integrated<br>gradients |  |  | - | 5.737e-02 | 4.695e-07 | 9.395e-07 | 2.331e-06 |
| random |  |  |  | - | 5.747e-10 | 9.201e-10 | 1.751e-09 |
| rise<br>(binomial) |  |  |  |  | - | 1.0 | 2.214e-01 |
| rise<br>(uniform) |  |  |  |  |  | - | 1.0 |
| shapley |  |  |  |  |  |  | - |
| Deletion: Mean over per whole slide image averaged AUC |  |  |  |  |  |  |  |
| Attn. Roll.<br>(w/o res) | - | 2.706e-01 | 4.647e-02 | 1.470e-07 | 7.828e-10 | 1.295e-07 | 6.089e-13 |
| Attn. Roll.<br>(with res) |  | - | 3.344e-05 | 6.745e-12 | 7.535e-06 | 4.610e-04 | 1.186e-08 |
| integrated<br>gradients |  |  | - | 1.030e-02 | 1.377e-15 | 5.454e-13 | 7.011e-19 |
| random |  |  |  | - | 1.716e-23 | 5.881e-20 | 1.975e-28 |
| rise<br>(binomial) |  |  |  |  | - | 1.0 | 1.0 |
| rise<br>(uniform) |  |  |  |  |  | - | 3.034e-01 |
| shapley |  |  |  |  |  |  | - |
| Insertion: Mean over all individual patch AUCs |  |  |  |  |  |  |  |
| Attn. Roll.<br>(w/o res) | - | 2.391e-40 | 1.419e-34 | 1.654e-36 | 0.0 | 0.0 | 0.0 |
| Attn. Roll.<br>(with res) |  | - | 1.0 | 1.321e-143 | 0.0 | 0.0 | 0.0 |
| integrated<br>gradients |  |  | - | 7.028e-133 | 0.0 | 0.0 | 0.0 |
| random |  |  |  | - | 0.0 | 0.0 | 0.0 |
| rise<br>(binomial) |  |  |  |  | - | 6.872e-08 | 8.071e-30 |
| rise<br>(uniform) |  |  |  |  |  | - | 1.332e-07 |
| shapley |  |  |  |  |  |  | - |
| Deletion: Mean over over all individual patch AUCs |  |  |  |  |  |  |  |
| Attn. Roll.<br>(w/o res) | - | 6.871e-66 | 2.724e-180 | 0.0 | 0.0 | 0.0 | 0.0 |
| Attn. Roll.<br>(with res) |  | - | 0.0 | 0.0 | 0.0 | 0.0 | 0.0 |
| integrated<br>gradients |  |  | - | 9.048e-159 | 0.0 | 0.0 | 0.0 |
| random |  |  |  | - | 0.0 | 0.0 | 0.0 |
| rise<br>(binomial) |  |  |  |  | - | 1.290e-34 | 7.128e-47 |
| rise<br>(uniform) |  |  |  |  |  | - | 1.698e-160 |
| shapley |  |  |  |  |  |  | - |

## Data Availability

All data produced are available online at https://camelyon16.grand-challenge.org/Data/.

## Data Availability

We leveraged the publicly available CAMELYON16 data set which can be found at https://camelyon16.grand-challenge.org/Data/.

## Code Availability

Code will be made available upon acceptance.

## Acknowledgements

We would like to thank Prof. Thomas Huegle from CHUV Lausanne for his valuable clinical advice. This project was supported by the Mertelsmann Foundation.

## Author contributions statement

JR, MN, GK and MK conceived the study. JR and MN conducted the experiments and performed the quantitative analysis. ED conducted the qualitative assessment. JB and MK provided strategic advice. JR, MK and GK wrote the manuscript. All authors approved the final manuscript.

## Footnotes

1 Due to computational limitations and the noisy nature of the Faithfulness metric, we omit this metric for the *Camelyon16* dataset.

2 https://timm.fast.ai/

